# From Tumor Board Presence to Documented Review: A Coverage Cascade for Multidisciplinary Cancer Care in India

**DOI:** 10.64898/2026.10.01.26364522

**Authors:** Vishala Bodetti, Nrip Nihalani, Rashmi Nivsarkar, Aditya Patkar

## Abstract

**Background:** Multidisciplinary tumor boards (MTBs) are a standard part of cancer care, yet India has no national measure of how many cancer patients are discussed at one. A national survey of hospitals in the National Cancer Registry Programme (NCRP), published in 2026, gives the first structured look at board presence and functioning.

**Objective:** To describe how far hospital-level tumor board presence translates into all-case review, documentation and follow-up, and to define the data needed to measure patient-level coverage.

**Methods:** Secondary analysis of published aggregate data: the ICMR-NCDIR survey of 229 hospitals in the NCRP hospital-based cancer registry (HBCR) network (October 2024), the HBCR 2021 report, national incidence estimates (ICMR-NCDIR 2022; GLOBOCAN 2024) and two published single-center MTB audits. We calculated proportions with Wilson 95% confidence intervals, non-response bounds across all 229 hospitals, and logical bounds for joint conditions. No individual-level data were used.

**Results:** A total of 229 hospitals were invited and 172 responded (75.1%). Among the respondents, 137 (79.7%; 59.8% to 84.7% across all hospitals) reported a functional tumor board. Only 65 respondents (37.8%; 28.4% to 53.3%) reported boards that review all cases, 71 (41.3%) had a follow-up system, 50 (29.1%) used any electronic documentation and 7 (4.1%) always took part in cross-hospital reviews. The share of hospitals with both all-case review and a follow-up system could lie anywhere from 0% to 37.8% of respondents. Boards were reported more often in private (86.6%) than in public hospitals (72.5%). Two single-center audits reported about 800 case discussions (573 new patients) and 1,260 case discussions (about 917 patients) a year, but neither reported new registrations at the same hospital, so coverage could not be calculated.

**Conclusions:** Tumor board presence is common in India, but evidence of all-case review, documentation and follow-up is much thinner, and the share of patients reviewed nationally is unknown. We propose six indicators that hospital-based registries could report to measure coverage directly.

## Introduction

Cancer is a major public health problem in India. The National Cancer Registry Programme estimated 1,461,427 new cases in 2022 [1], and the GLOBOCAN 2024 estimate for India is 1,562,581 cases [2]. Diagnosis, staging and treatment planning involve several disciplines, including surgery, medical and radiation oncology, radiology and pathology. Multidisciplinary tumor board review is the usual way these disciplines coordinate. A systematic review of the oncology literature found that multidisciplinary team meetings affect how patients are assessed and managed, although it found limited evidence that they improve clinical outcomes such as survival [3].

Despite this, little is known about how widely tumor boards operate in India or what share of patients they reach. In 2026, ICMR-NCDIR published a national survey of hospitals registered under the NCRP hospital-based cancer registry network. It found that most responding hospitals had a functioning tumor board, but that documentation, follow-up and cross-hospital collaboration were limited [4]. Single-center audits show what a functioning board can handle. For example, a private comprehensive cancer hospital in Eastern India reported 800 case discussions (573 new patients) in one year [6], and a public tertiary hospital in Northeast India reported 4,704 case discussions (3,426 patients) over almost four years [7].

These sources describe different things. A hospital can have a board without discussing every patient, discuss patients without recording the outcome, or record the outcome without checking whether it was followed. Presence, coverage, documentation and follow-up are separate steps, and a survey of hospitals cannot say how many patients pass through each. This matters because planning and audit need a patient-level denominator: the share of new cancer patients who were discussed before treatment.

### Objectives

We aimed to (1) quantify, with explicit uncertainty, how far reported tumor board presence translates into all-case review, documentation and follow-up across hospitals in the NCRP network; (2) show what published single-center audits can and cannot tell us about patient-level coverage; and (3) propose the minimum data needed to measure coverage directly. This is a descriptive analysis of published aggregate data. It does not estimate the proportion of Indian patients who receive multidisciplinary review.

## Methods

### Design and data sources

We carried out a secondary analysis of published aggregate data from four sources. First, the ICMR-NCDIR cross-sectional survey of tumor boards in hospitals registered under the NCRP hospital-based cancer registry network, conducted from 1 to 31 October 2024 and published on 12 March 2026 [4]. Second, the ICMR-NCDIR report of the hospital-based cancer registries for 2021 [5]. Third, national cancer incidence estimates for 2022 from ICMR-NCDIR [1] and for 2024 from GLOBOCAN [2]. Fourth, two published single-center audits of tumor board activity in India that reported the number of cases discussed [6,7]. These were identified through a targeted literature search (not a systematic review), and we read both in full. We extracted only values stated in these publications. The number of hospitals with any electronic documentation was calculated as the sum of two reported categories: electronic only (n = 16) and both electronic and physical (n = 34).

### Derived quantities

For each survey indicator we calculated the proportion among respondents with a Wilson 95% confidence interval. Because 57 of 229 hospitals did not respond, we also calculated non-response bounds across all 229 hospitals: the lower bound assumes no non-respondent has the attribute, and the upper bound assumes every non-respondent has it. The source reports follow-up, documentation and cross-hospital review for the 137 hospitals with a board, so hospitals without a board were counted as not having these attributes. The survey does not report how attributes overlap within hospitals, so for hospitals that both review all cases and have a follow-up system we calculated logical (Fréchet) bounds from the two marginal counts. For the audits, we divided reported cases by the audit period to obtain annual throughput. We did not annualize the HBCR report’s case totals, because hospitals contributed different numbers of years.

### Ethics

Only published aggregate data were used. No individual-level data were collected, and ethics committee review was not required.

## Results

### Hospital frame and board characteristics

The NCRP network survey invited 229 hospitals, and 172 responded (75.1%). Half were publicly funded (52.3%). Tertiary-level centers (45.3%) and medical colleges (43.6%) made up most respondents. A total of 137 respondents (79.7%) reported having a functional tumor board. Of these boards, 71.5% were single boards, 73.7% met in person and 47.4% met weekly. Table 1 summarizes the characteristics reported in the source.

**Table 1.** Characteristics of hospitals and tumor boards in the NCRP hospital-based cancer registry survey (October 2024), as reported in [4].

| Characteristic | n | % (as reported) |
| --- | --- | --- |
| Hospitals registered under the network | 229 |  |
| Hospitals that responded | 172 | 75.1% |
| Public / private funding (among respondents) | 90 / 82 | 52.3% / 47.7% |
| Tertiary centers / medical colleges / regional cancer centers / secondary hospitals | 78 / 75 / 17 / 2 | 45.3% / 43.6% / 9.8% / 1.2% |
| Hospitals with a functional tumor board | 137 | 79.7% |
| Board present: private vs public hospitals | — | 86.6% vs 72.5% ($\chi^2 = 5.17$, $p = 0.023$, as reported) |
| Hospitals with multiple boards (among boards) | 39 | 28.5% |
| In-person / hybrid / fully virtual meetings | 101 / 34 / 2 | 73.7% / 24.8% / 1.5% |
| Weekly meetings | 65 | 47.4% |
| As-needed or irregular meetings | 39 | 28.5% |
| Annual meeting calendar maintained | 87 | 63.5% |
| Designated secretariat | 72 | 52.5% |
| Review all cases treated / specific cases only | 65 / 72 | 47.5% / 52.5% |
| Documentation method: electronic / physical / both | 16 / 87 / 34 | 11.7% / 63.5% / 24.8% |
| Recommendations communicated by EMR notes / written summaries / direct contact / other | 23 / 52 / 48 / 14 | 16.8% / 38.0% / 35.0% / 10.2% |
| Follow-up system for board recommendations | 71 | 51.8% |
| Recommendations implemented “most of the time” / “always” | 81 / 40 | 59.1% / 29.2% |
| Cross-hospital case discussion: always / sometimes / never | 7 / 49 / 81 | 5.1% / 35.8% / 59.1% |
| Participation: radiation oncologists / surgical oncologists / medical oncologists | — | 92.7% / 83.2% / 78.8% |

### The coverage cascade

Among all 172 respondents, 79.7% (95% CI 73.0–85.0) reported a board, but fewer reported the features needed for patient-level coverage (Table 2, Figure 1). Boards that review all cases treated at the hospital were reported by 65 respondents (37.8%; 95% CI 30.9–45.2). A follow-up system was reported by 71 (41.3%; 34.2–48.7), any electronic documentation by 50 (29.1%; 22.8–36.3) and cross-hospital case discussion in every case by 7 (4.1%; 2.0–8.2). A further 49 hospitals reported cross-hospital case discussion only sometimes. If no non-responding hospital had these features, the corresponding shares across all 229 hospitals would be 59.8%, 28.4%, 31.0%, 21.8% and 3.1%. If every non-responding hospital had them, the shares would be 84.7%, 53.3%, 55.9%, 46.7% and 27.9%.

**Table 2.**
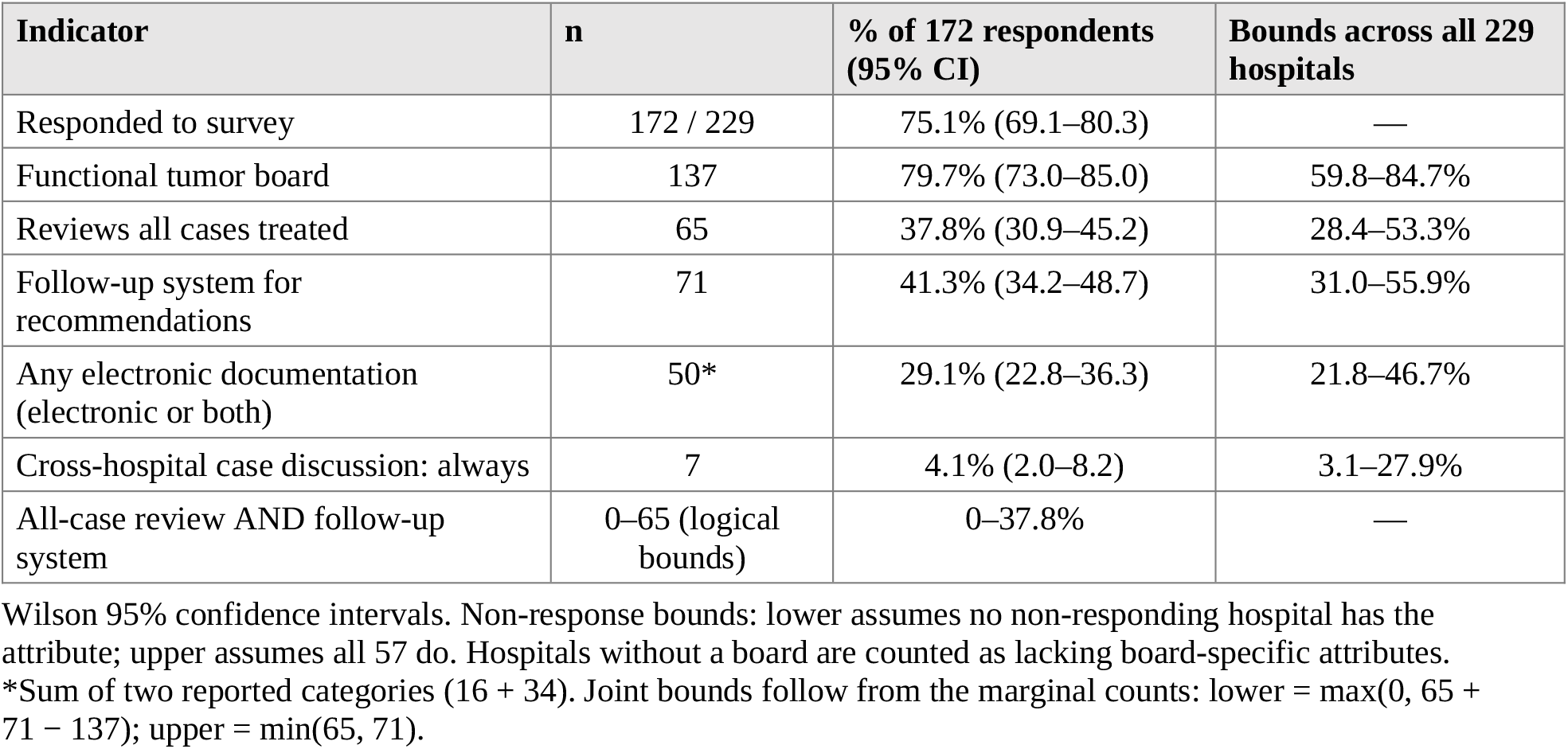
Hospital-level coverage cascade with sampling and non-response uncertainty.

**Figure 1.**
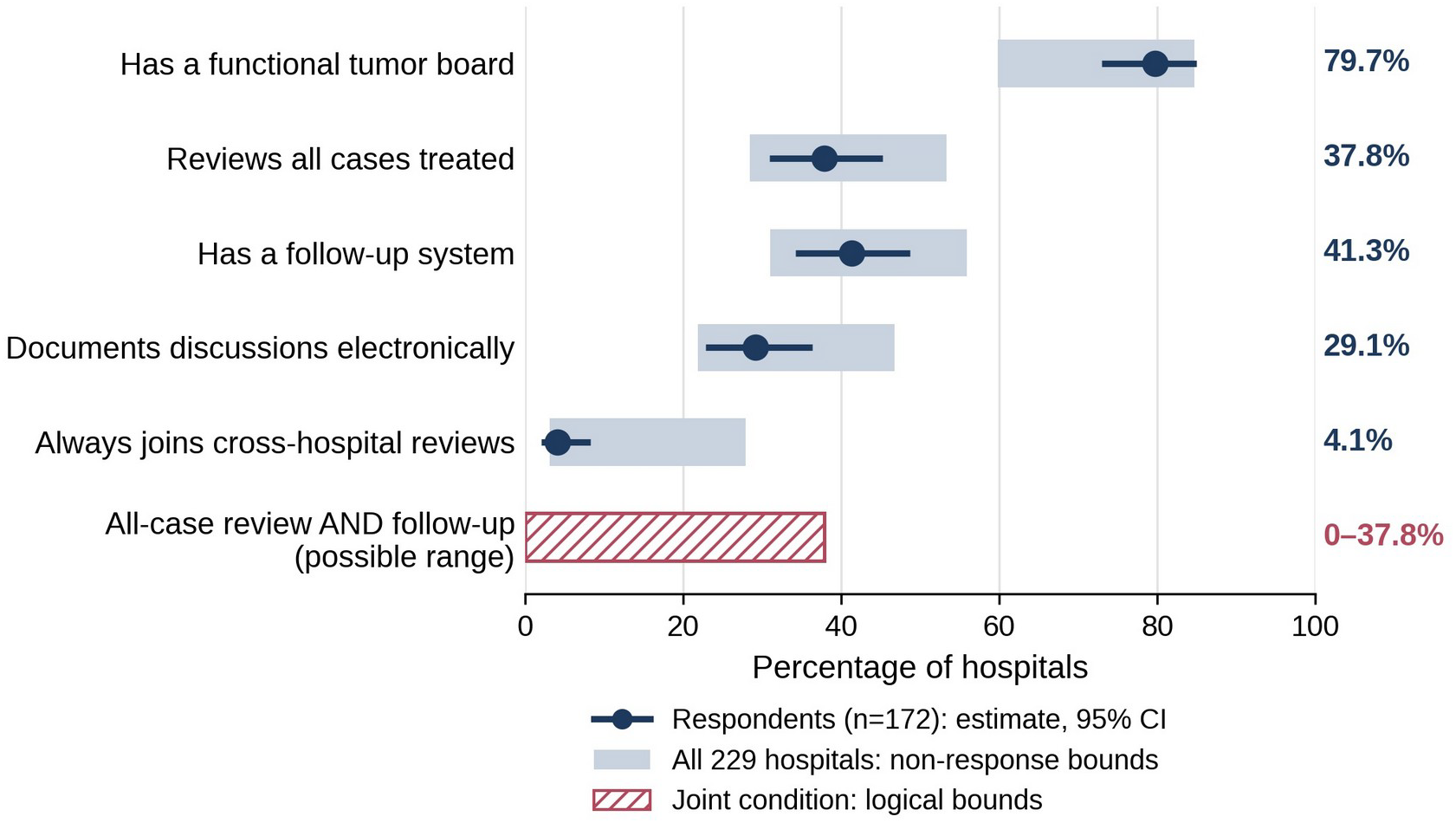
Hospital-level coverage cascade. Dots with lines show the estimate and 95% confidence interval among 172 respondents; grey bars show bounds across all 229 hospitals if non-respondents all lacked or all had the attribute; the hatched bar shows the logical range for hospitals with both all-case review and a follow-up system.

The survey does not report how all-case review and follow-up overlap. Because 65 hospitals review all cases and 71 have follow-up systems, and both groups are contained within the 137 hospitals with a board, the number meeting both conditions could be anywhere from 0 to 65 hospitals, or 0% to 37.8% of respondents. The published data cannot narrow this range.

### Public and private hospitals

The source reported that boards were more common in private (86.6%) than in public hospitals (72.5%) (χ^2^ = 5.17, p = 0.023) [4]. Counts for the public group were not given. When we tried to reproduce the test from the reported percentages and group sizes, we obtained similar but not identical statistics (χ^2^ between 4.65 and 5.35, depending on the rounding of the public count), so we report the source figures as stated.

### Documented activity at audited boards

Two published audits reported the volume of cases discussed at a single hospital (Table 3). The Northeast India board discussed 4,704 cases (3,426 patients) between 14 June 2016 and 9 March 2020 [7], which is about 1,260 case discussions and 917 patients per year over 3.73 years. The Eastern India board discussed 800 cases in a 12-month period, of which 573 were new cases and 227 were repeat discussions [6]. That paper gives the period as September 2020 to August 2021 in its abstract and August 2020 to July 2021 in its Methods. The Eastern India board met once a week and the Northeast India board twice a week. We read both audits in full: neither reports the number of new cancer registrations at the same hospital, so neither can show what share of new patients were discussed. The two audits also count different units (case discussions versus patients).

**Table 3.**
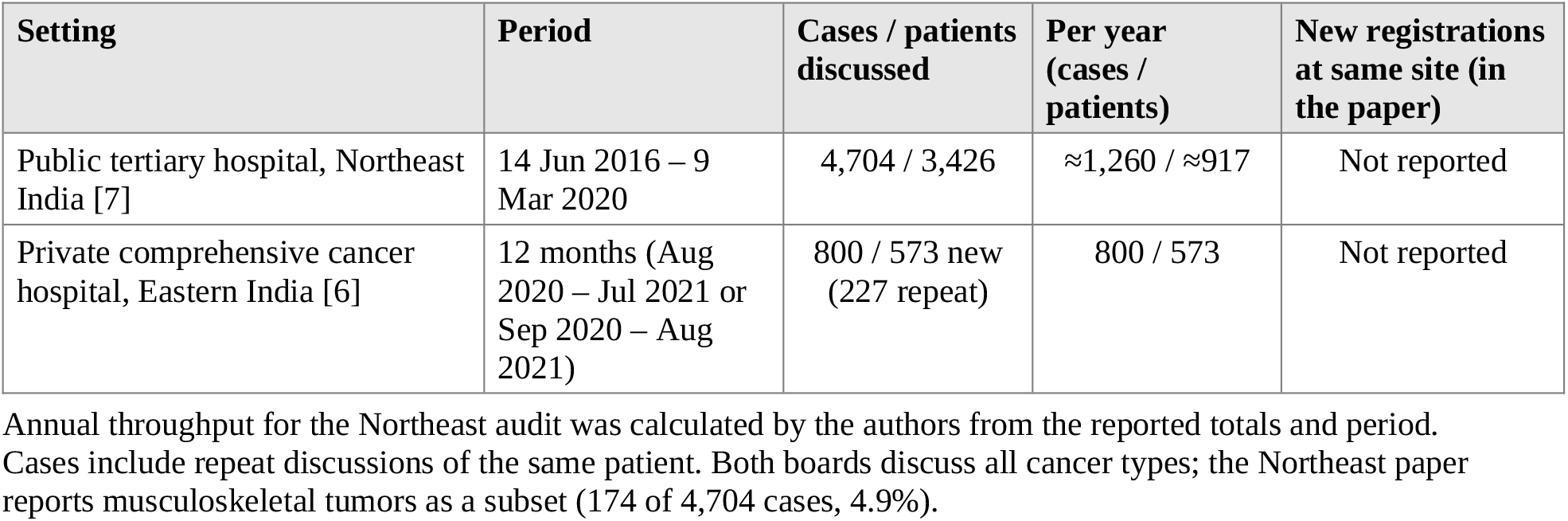
Published single-center tumor board audits from India.

### Scale context

The HBCR 2021 report registered 1,332,207 new cancer cases from 96 hospitals during 2012–19, and 610,084 of these had complete details and were analyzed [5]. The 96 hospitals were selected from 269 registered HBCRs because they had completed data transmission and quality checks for one or more years, so they contributed different numbers of years and an annual volume cannot be derived reliably from the totals. The 2024 survey frame of 229 hospitals is also different from the report’s 96. The report therefore shows that registry hospitals hold large case volumes, but it does not provide hospital-level annual counts that could weight the survey results. Without such counts, hospital-level presence of a board cannot be translated into national patient coverage.

## Discussion

We found that tumor board presence is common among hospitals in the national registry network, but that the features needed to show patient-level coverage are far less common. About four in five responding hospitals reported a board, fewer than two in five reported a board that reviews all cases, and about two in five had a follow-up system. Fewer than one in three used any electronic documentation, and only a small minority always took part in cross-hospital reviews. Even under the most favourable assumption about non-responders, fewer than 56% of hospitals would have a follow-up system or all-case review. The proportion with both could not be determined from the published data.

The main finding is a measurement gap. National incidence is well estimated [1,2], and hospitals report whether a board exists [4], but no source reports how many new patients were discussed before treatment. The audits show that individual boards can handle roughly 800 to 1,260 case discussions a year, equal to about 573 to 917 patients [6,7], but without a hospital’s new registrations, they do not show what proportion of patients that represents. This gap limits planning, audit and quality improvement. It also means that statements about the reach of multidisciplinary care, from any organization, cannot be checked against national data.

Two other findings from the source survey deserve attention. First, boards were reported more often in private than in public hospitals [4], which, if confirmed, points to an access difference by sector. Second, cross-hospital case discussion was reported as always by 5.1% and as sometimes by 35.8% [4], so boards at smaller hospitals may lack routine specialist input that virtual or hybrid formats could provide.

The registries already collect diagnosis, stage and treatment details [4]. Adding a small number of fields would allow coverage to be measured directly. We propose six indicators (Table 4). Each uses a numerator and denominator that a hospital-based registry could report, so that coverage can be compared across hospitals and over time.

**Table 4.** Proposed minimum indicators for measuring tumor board coverage (authors’ proposal).

| Indicator | Numerator / measure | Denominator |
| --- | --- | --- |
| 1. Case coverage | New analytic cases discussed at a board before first definitive treatment | New analytic cases registered |
| 2. Timeliness | Median days from registration to board discussion | — |
| 3. Documentation | Discussed cases with a written, retrievable recommendation in the record | Cases discussed |
| 4. Concordance | Recommendations implemented as advised, or with a documented reason for deviation | Cases with a recommendation |
| 5. Management change | Discussed cases with a change in diagnosis, stage or plan | Cases discussed |
| 6. Cross-site reach | Cases discussed through cross-hospital or virtual boards | Cases discussed |

### Strengths and limitations

Strengths are the use of a recent national source, explicit uncertainty (confidence intervals and non-response bounds) and a clear statement of what the data cannot show. The study has limitations. The survey counts hospitals rather than patients, the data are self-reported and were collected over one month, and 25% of invited hospitals did not respond. The survey did not report how attributes overlap, so joint conditions are given as bounds. We could not access hospital-level data, which would allow weighting by patient volume; a data request to the registry programme would be the natural next step. The audits were identified through a targeted rather than systematic search, and one of them gives two different 12-month periods in its abstract and Methods. The HBCR report covers 2012–19 and hospitals contributed different numbers of years, so we did not annualize its volume.

We noted internal inconsistencies in the source survey’s summary figures. The share of hospitals with a single board is 71.5% in the Results and 75.1% in the Discussion. Electronic records are given as 26.3% in the Discussion, whereas the documentation table implies 36.5% (16 electronic plus 34 both). Public-hospital counts for the board comparison are not reported. We used the counts in the source tables throughout. Finally, the proposed indicators are untested.

## Conclusions

Tumor board presence is common in India’s registry hospitals, but evidence of all-case review, documentation and follow-up is much thinner, and the share of patients reviewed nationally is unknown. Measuring coverage requires a patient-level denominator. Adding a small set of indicators to hospital-based cancer registries would allow it.

## Declarations

### Ethics approval

Not applicable. The study used only published aggregate data and involved no human participants or individual-level data.

### Author contributions

Dr. Vishala Bodetti carried out the main research and the documentation. Nrip Nihalani, Dr. Rashmi Nivsarkar and Aditya Patkar reviewed the manuscript. All authors read and approved the final version.

### Declaration of generative AI use

During the preparation of this manuscript, the authors used AI tools to edit the text, to help design the tables and figures, and to assist with identifying published sources. AI tools are not authors. The authors reviewed and verified all analysis and take full responsibility for the content, accuracy, and integrity of this work.

### Data availability

All data are from the cited public sources. The derived calculations are described in the Methods and Tables 1–3 and can be reproduced from the reported counts.

